# Low COVID-19 mortality in old age homes in western India: an empirical study

**DOI:** 10.1101/2020.12.08.20245134

**Authors:** Jallavi Panchamia, Bhavya Bhagat, Vishakha Bharati, Anushree Joshi, Dileep Mavalankar

**Affiliations:** Indian Institute of Public Health Gandhinagar, India

**Keywords:** Old age homes, COVID-19, low mortality

## Abstract

**Objective:** The study intended to understand the effect of Coronavirus disease 2019 (COVID-19) on western Indian elderly care homes. The study aimed to know the actions taken by administration of homes and challenges faced during the lockdown period.

**Method:** Total 44 care homes across three states of western India were contacted for data collection during the period of September-November 2020. Structured interview of the manager or owner of the elderly care homes were taken to gather required information to achieve the study objectives.

**Results:** Out of 44 care homes, seven care homes reported 146 case of corona virus infection and four deaths. Hence, the reported covid cases rate found to be 928 per 10,000 residents.

**Discussion:** Results of the study indicated that significant steps were taken by these old age care homes to stave off the infection spread among the inmates. It was observed that average 26% of the occupants were sent back to their home/relative’s home before the lockdown to decongest the care homes. Administrators adopted the new policies at care home and adhered the government guidelines. Care homes in western India seemed to have very low infection rate and very low number of deaths as compared to western world.

## 1. Background

The elderlies are by far the most vulnerable population group in context to prone to infection of COVID-19 disease. As per the reports, age is the most important determinant in magnitude of the spread and effect of COVID-19 disease especially after 65 years of age (World Health Organization, 2020). It has become significant to take extra care for elderly person at home, but when it comes to old age homes, this becomes a huge challenge since not just one or two, all the occupants need special care to avoid getting infected. As care homes have a unique, mixed population of multi-disciplinary staff and frail residents with multiple underlying comorbidities, it becomes difficult to avoid any occurrence of infection spread. Elderly residents are at high risk of severe complications and death due to respiratory viruses, such as influenza etc. including coronavirus.

During the initial months of COVID pandemic, community care facilities including nursing and residential homes were termed as “hubs” and “besieged castles” in North America and Europe^1^, having experienced large outbreaks due to rapid transmission of SARS-CoV-2. whereas, Canada experienced high number of coronavirus-related deaths in elderly care homes, according to a Canadian Institute for Health Information report. As per the global reports, England and Wales observed 45,899 deaths among care home residents between March 02 and May 02 and 12,526 got infected by COVID-19^1^. Similar experiences have been observed in France, Spain and the United States, where only limited measures were taken. But conversely countries like Australia, Austria and Slovenia, which implemented “specific prevention measures” targeted at elderly care homes, including segregating units and wide screening for the illness, have reported fewer COVID-19 infections and deaths (“Canada ranks worst in elderly care home coronavirus deaths: study”, 2020).

Meanwhile, in India as cases are rising abruptly, deaths are recorded on daily basis. Death rate among elderly is high in India. On other hand, hardly any studies have reported any data on the numbers of elderly residents of long-term care facilities such as nursing homes, old age homes, who might have been infected or died from confirmed or suspected coronavirus. Hence, the present study intended to understand the effect of Coronavirus disease 2019 (COVID-19) on Indian elderly care homes. The study also aimed to know the actions taken by administration of homes and challenges faced during the lockdown period. It would be noteworthy to understand the managerial actions taken by Indian elderly care home during such difficult time of pandemic.

### 2. Literature Review

Since last few years, old age homes across the nations have increased in number due to urbanization, migration, frequent relocating due to job opportunities, Increased living costs, internal disputes between close relatives, and life style changes (van Bilsen et al., 2006). As the demand of care homes have increased, numerous challenges too have grown. As some of the elderly might be suffering from debilitating diseases and can be on bed rest, management needs to train the special staff to take care of them (Zhang et al., 2020). One of the ongoing challenges is higher intake of residents then capacity which becomes difficult for old age homes to sustain the expenses. Sourcing funds becomes the one of the challenges faced by old age homes when they are working on trust-based or private funding. From the occupants’ side, old age care homes sometimes fail to take care of the elderly properly. Moreover, rules are too strict in terms of visiting their loved ones, loneliness and sometimes abuse and neglect, are the main issues faced by them (Gelfand, 1968). According to the study, Old people who live in family settings have higher efficiency levels and social interactions (Dubey et al., 2011).

COVID-19 has taken the world by storm. According to the World Health Organization, elderly aged more than 50 years are highly susceptible to developing severe complications of COVID-19 (Older People and COVID-19, n.d.). As elderlies are the age group where if the care is not taken, even due to the aging factors body’s immunity goes down. Which makes them one of the most susceptible group for encountering diseases frequently (Asadullah et al., 2012). Hence, it is widely practiced to keep elderly people in a separate space or isolation due to the fear of family members transmitting infection to them and others. This has resulted into higher level of anxiety and loneliness among elderly (Tiwari et al., 2012).

Few studies from USA, Canada and UK highlighted the difficulties faced by elderly care homes amidst pandemic such as shortage of personal protecting kits and care giver staff, less preparedness of management to face the surge of cases and higher mortality rate among elderly (Gordon et al., 2020; McMichael et al., 2020; Ladhani et al., 2020; Comas-Herrera et al.,2020). Some of the care homes in UK had staff serving multiple facilities which made them a carrier and caused the spread of infection from one place to another (McMichael et al., 2020). The earlier guidelines of testing only those patients who have symptoms is not effective as in London care homes 49% of those harbouring infection were asymptomatic (Ladhani et al., 2020).

In India, according to the grey literature and news reports published, the southern part of India reported COVID-19 cases inside the old age home (Care Homes on High Alert-The New Indian Express, n.d.) from Kerala, Tamil nadu and Karnataka, whereas few inmates diagnosed with COVID-19 in Navi-Mumbai during the month of October-November,2020(Shaju Philip, 2020, Vijay Kumar, S, 2020, Theresa, D, 2020). It is surprising to note that only few news reports documented the status of old age care homes in other states of India.

The study was conducted to explore the situation in few states of India such as Gujarat, Maharashtra, and Rajasthan, where higher number of covid infection cases are substantial in general. Although, none of the study focused to the situation of covid in old age home.

### 3. Research Design

Data was collected by trained doctoral scholar students, Research associates and faculty through telephonic interview method. Respondents in this study were mainly administrators/ owner/ manager of care home in three states namely, Gujarat, Maharashtra and Rajasthan. These three states were selected based on two rationale, first being states with high rate of mortality and covid infection case reported and second being the ease of reaching to the care home respondents due to available contacts and references. To allow for heterogeneous sample inclusion, respondents from care homes with different ownership (public/trust based and private) and geographic locations (rural and urban) were contacted for data collection. Data collection was conducted through telephonic survey, since it was not feasible to travel in wake of government guidelines for physical distancing and safety.

Total 52 care homes were contacted during the months of September-November 2020, out of which 44 responded. They gave consent to give macro level information such as current status of covid cases/deaths, demographic profile of occupants and related statistics before and after lockdown, facilities, specific precautions due to pandemic, challenges, and policy changes in wake of such difficult time.

Data collection was done by using semi-structured interviews, with each interview lasting for about 20 minutes. Initial questions included information like name and address of care homes, followed by details such as rate of occupancy before and during pandemic, reported/suspected covid infection case and other related information. Information related to individual occupants of care home were not taken since it was not required as per the scope of the study. Although, information related to demographic composition in terms of gender, age and co-morbidities of occupants was collected. Moreover, Care homes’ responses to pandemic were captured by asking the open-ended questions related to challenges faced by administrators, available resources/ facilities and actions taken to avoid occurrence and transmission of infection among elderly residents. These questions were followed by questions related to their resourcefulness, staff and medical treatment accessibility and availability; for instance, frequency of doctors/nurses’ visits, isolation or quarantine facility, and basic equipment/medical kit such as thermometer, Blood-pressure measuring equipment, Oximeter and Oxygen cylinder. Subsequently, probing questions related to staff shortage during lockdown followed.

Right at the outset, the participants were informed about the objectives of the study, and explicit verbal consent was obtained for conducting the interviews. Additionally, the authors gave assurance to the respondents that the interviews were solely for the purpose of research and was not a sponsored exercise to evaluate the care homes objectively; anonymity was also assured and safeguarded in the analysis.

### 4. Findings

Key findings are given in the table 1 below.

**Table 1:** Characteristics of elderly care homes included in the study (n= 44)

|  | Gujarat | Maharashtra | Rajasthan | total |
| --- | --- | --- | --- | --- |
| Total old age home surveyed in India | 13 | 17 | 14 | 44 |
| Types of ownership |  |  |  |  |
| Private | 7 | 10 | 7 | 24 |
| Trust based/Public | 6 | 7 | 7 | 20 |
| Occupancy before lockdown (pre-COVID) | 680 | 820 | 614 | 2114 |
| Occupancy during COVID pandemic | 525 (77 %) | 598 (72 %) | 450 (73 %) | 1573(74%) |
| Occupants-Gender wise distribution |  |  |  |  |
| Male | 40% | 71% | 47% | 51% |
| Female | 60% | 29% | 52% | 49% |
| COVID cases reported | 41 | 71 | 34 | 146 |
| Deaths reported due to COVID | 2 | 1 | 1 | 4 |

Maharashtra (total cases = 18,47,509, & 16.180 cases per million), Gujarat (total cases = 2,17,333, & 3466 cases per million) and Rajasthan (total cases = 2,78,496 & 4043 cases per million) are the states situated in the western part of India and showing the higher number of cases in comparison to the other states. The most affected districts are Mumbai (total cases = 2,79,737,& 15,203 cases per million), Pune (total cases = 168866, & 54,123 cases per million) & Nagpur (total cases=24,10,000, & 48,180 cases per million) in Maharashtra, Ahmedabad (total cases = 47,757 & 8573 cases per million), Rajkot (total cases = 10,019, & 777 cases per million) & Surat (total cases = 38,472, & 863 cases per million) in Gujarat and Jaipur (total cases = 48,463,& 15,786 cases per million) in Rajasthan as of 6th December, 2020. The old age care homes included in the study are mainly situation in and around these districts.

As we can see from the summary table, three care homes out of 13 reported 41 cases and two deaths in Gujarat, four care homes out of 17 reported 71 cases and one death in Maharashtra, whereas three old age homes out of 14 reported 34 cases and one death in Rajasthan till we did the inquiry, i.e. 8^th^ December, 2020. It was found from these survey that average 26 % of the elderly residents left the home before the lockdown, i.e. during March end. This thing was seen in total 39 old age homes across the three states, where residents were sent back.

## 5. Discussion

### Reported SARS COV-2 cases in care homes

One of the old age homes in Gujarat reported 35 cases with the transmission from the administration department employee who found to be asymptomatic covid infected. As per the information shared by the administrator, they all were shifted to covid care centre and all recovered. Another care home resident was diagnosed of covid when he was undergoing thru dialysis in the hospital. He died in the hospital itself and not in the old age home. One of the care homes in Maharashtra reported 19 covid cases during the last week of November out of the total residents of 60, whereas another care home near Mumbai reported 40 cases and no death. These care homes surprisingly reported no death in spite of significant number of cases.

One of the covid case, which happened in the month of April in Gujarat, was detected during the government screening and appeared to be asymptomatic. The patient in her 60’s was admitted in civil hospital for 25 days as gradually symptoms were developed and she returned to the old age home after she tested negative. One of the old age homes near Jaipur in Rajasthan reported 25 cases out of the occupancy of 100 and did not report a single death.

It was found that 40 old age homes out of 44 did not have any cases till August, 2020. Although later more than 100 cases were reported in these surveyed homes during the period of September-November, only four deaths were reported. These findings contradict the situation of the old age care homes in other countries, where majority of the cases and deaths reported from old age homes (Gordon et al., 2020; McMichael et al., 2020; Ladhani et al., 2020; Comas-Herrera et al.,2020).

### Resourcefulness of the care homes

It was found that 100 % care homes were equipped with thermometer, 95 % with blood pressure measuring equipment, 70 % with Oximeter but only 5 % with oxygen cylinder. All the care homes had regular visit of doctors, although frequency was varying from weekly to monthly. In fact, 3 care homes had a facility of resident doctors for medical emergency and well-equipped hospitals to refer. While 2 care homes had a difficulty reaching out to hospitals during referral and the availability of ICU beds for COVID patients. 80 % of the care homes had a facility of isolation and quarantine ready if any cases arise.

Only 18 % of the care homes faced the staff shortage and higher absenteeism during and after lockdown period mainly because of their locational disadvantage being far from the city and lack of residential area nearby.

85% of the care homes were adequately funded and had no issues with resource allocation to meet needs of residents, but in 15 % of the care homes, the situation was the opposite as resources were scanty to meet the requirement. In some of such NGO-trust based care homes, trustee kept the homes in good shape by investing their personal money as a philanthropic activity.

### Precautions and steps taken to avoid getting infection

The administrators from care homes shared the actions taken by them to keep safe their elderly residents from getting infected of corona virus. The home administrators first stopped all contact with outside world as much as possible. To stop infection getting spread in the old age homes, the administrators ensured that the staff were made to live temporarily in the old age homes, thus preventing contact with outside world.

Interestingly, 65 % of the care homes insisted their staff to stay in care homes and not commute daily from their own homes. Rest of the care homes had strict policy for their staff to get sanitised before entering into the premise of care homes. This was not seen in majority of the western care homes, where rotation of staff among care homes are practiced (McMichael et al., 2020). Literature pointed out that infection among care home residents was very high in western country, because staff were moving around after their shift used to get over in care home, eventfully they got infected and passed it to elderly residents (Padmanabha et al., 2016). Hence, such step of insisting the old age home staff to either stay in the care homes or entry after through sanitisation helped in stopping the spread of infection.

6 Old age homes from Gujarat, 10 from Maharashtra and 5 from Rajasthan saw decrease in elderly population due to sending them back to their relatives to decongest the Old age homes Thus, it was somewhat easy for them to manage the smaller number of residents and following the physical distancing norms from all aspects.

They ensured proper sanitization of all the outside items such as vegetables etc before entering in the premise. They also took utmost care of proper sanitization of the old age home premise on a frequent basis. All elderly residents, caretakers, staff and administrative staff were educated for maintaining social distancing wearing masks, sanitizing, other safety protocols, and preparedness. Most of the respondents from care homes confirmed that the safety protocols of fumigation and deep sanitation were taken periodically.

Most of the care home administrators brought dietary changes and added certain physical activity to the routine of residents to keep them engaged and healthy. In order to boost he immunity among them, vitamin. C, D, and multivitamins and ayurvedic “*kadha*” were added to their diet. The elderlies were encouraged to make video calls to their families and showing them movies and ensuring a secure space by interacting environment to reduce their anxiety from the chaos. No inmate was allowed to go outside and there was no contact between staff which use to go outside to procure resources. In fact, two of the care homes took the special care for their bedridden elderly to avoid an outbreak situation.

### Challenges faced by Old age homes

It was challenging for some old age care homes which were managed on a very shoe string budgets by non-professional philanthropic organization to ward off the probability of infection spread among their residents. Generally, they used to take their elderly residents for some half day or short excursion to nearby areas to bring some change in their monotonous daily routine, but due to such unprecedented pandemic time, they could not do so. Most of the care home administrators mentioned this situation as challenging. Moreover, they could not allow their family/friends also, who generally used to come to spend some time with their elderly residents, or for some donation and celebrations. Thus, this situation brought lot of dullness in the lives of residents.

One of the administrators from Nashik said during the interview “we have some inmates who are homeless and have no families to look after them. They are completely dependent on us, so their physical and mental well-being is our responsibility. I want to urge the authorities to regulate the news channels and other media on which all the time new will be broadcasted related to deaths due to COVID-19 and chaos in different parts of the world due to COVID-19 They could have shown favourable cases which have recovered and about the good recovery rate also along with that news so that it can bring some solace among these elderly people.”. Hence, this pandemic time has brought lot of anxiety and mental well-being issues among elderly residents staying in old age care homes away from their loved ones. Although, some of the old age home mangers made a provision for their residents of video call with families, some lifestyle changes such as practicing yoga and meditation etc. some of the administrators also arranged mental health counselling sessions for residents to improve the level of their psychological well-being. Few of the care homes had the enough fund to survive, whereas majority of the old age homes faced substantial decrease in the number of donors and amount funds.

## 6. Limitations of the study

Since the study was done through telephonic survey, there was an absence of non-verbal cues like observing their expressions, emotions with which they talk, observing the facility itself. Some of the respondents may not have given full information during the interview due to the fear of loss of image or lack of trust., hence, the authenticity of the responses was not confirmed. There might be some sort of respondent’s bias about giving positive or distorted answers regarding the covid situations to showcase the favourable picture. These gaps in the study need to be addressed with on-site observations. This study was conducted at macro level, which could not consider the perspectives and responses of old-age home residents. Findings could have been very different if study could have been conducted taking all the old age occupants as respondents. This study considered only tested and confirmed covid cases and not the suspected cases. Results could have been different if sero-surveillance would have been done with blood testing and our interviews could have been conducted earlier in the situation at the starting of lockdown period.

## 7. Conclusion

The notable point is most of the observations are similar in these three states of India. It was surprising to see very low mortality rate in old age home of all the three states. To stop infection getting spread in the old age homes, the managers ensured that the care workers were made to live temporarily in the old age homes, thus preventing contact with outside world. They ensured proper sanitization of all the outside items such as vegetables etc before entering in the premise. They also took utmost care of proper sanitization of the old age home premise on a frequent basis. Testing of inmates was done before taking new admission in few institutions and for rest, they remain closed for new entries to avoid the outbreak. In fact, they also sent back some of the elderly to their relatives’ home those who could in order to decongest the old age homes. It would be interesting for clinical researchers to understand the reason behind the low mortality and low infection rate in old age homes apart from the other reasons cited in the paper.

## Data Availability

data will be available on request.

## References

Bansal, R., Bansal, M., & Kumar, M. (2008). Need to support old-age home residents. Indian Journal of Community Medicine, 33(2), 131. https://doi.org/10.4103/0970-0218.40887

Booth, R. (2020, May 18). Agency staff were spreading Covid-19 between care homes, PHE found in April. The Guardian. https://www.theguardian.com/world/2020/may/18/agency-staff-were-spreading-covid-19-between-care-homes-phe-found-in-april

Canada ranks worst in elderly care home coronavirus deaths: study. (2020, June 25). Medicalxpress.https://medicalxpress.com/news/2020-06-canada-worst-elderly-home-coronavirus.html

Dubey, A., Bhasin, S., Gupta, N., & Sharma, N. (2011). A study of elderly living in old age home and within family set-up in Jammu. Studies on Home and Community Science, 5(2), 93–98. https://doi.org/10.1080/09737189.2011.11885333

Faghanipour, S., Monteverde, S., & Peter, E. (2020). COVID-19-related deaths in long-term care: The moral failure to care and prepare. Nursing Ethics, 27(5), 1171 –1173. https://doi.org/10.1177/0969733020939667

Gelfand, D. E. (1968). Visiting patterns and social adjustment in an old age home. The Gerontologist, 8(4), 272–304. https://doi.org/10.1093/geront/8.4.272

Gordon, A. L., Goodman, C., Achterberg, W., Barker, R. O., Burns, E., Hanratty, B., Martin, F. C., Meyer, J., O’Neill, D., Schols, J., & Spilsbury, K. (2020). Commentary: COVID in care homes—challenges and dilemmas in healthcare delivery. Age and Ageing, 49(5), 701–705. https://doi.org/10.1093/ageing/afaa113

Inmates of old age homes, orphanages being tested for COVID-19. (2020, July 31). THE HINDU. https://www.thehindu.com/news/cities/Tiruchirapalli/inmates-of-old-age-homes-orphanages-being-tested-for-covid-19/article32241467.ece

Ladhani, S., Chow, J. Y., Janarthanan, R., Fok, J., Crawley-Boevey, E., Vusirikala, A., Fernandez, E., Sanchez Perez, M., Tang, S., Dun-Campbell, K., Wynne-Evans, E., Bell, A., Patel, B., Amin-Chowdhury, Z., Aiano, F., Paranthaman, K., Ma, T., Saavedra-Campos, M., Myers, R., … Zambon, M. (2020). Investigation of SARS-Cov-2 outbreaks in six care homes in London, April 2020: The London care home investigation. SSRN Electronic Journal. https://doi.org/10.2139/ssrn.3638267

McMichael, T. M., Currie, D. W., Clark, S., Pogosjans, S., Kay, M., Schwartz, N. G., Lewis, J., Baer, A., Kawakami, V., Lukoff, M. D., Ferro, J., Brostrom-Smith, C., Rea, T. D., Sayre, M. R., Riedo, F. X., Russell, D., Hiatt, B., Montgomery, P., Rao, A. K., … Duchin, J. S. (2020). Epidemiology of COVID-19 in a long-term care facility in King County, Washington. New England Journal of Medicine, 382(21), 2005–2011. https://doi.org/10.1056/nejmoa2005412

Mendonca, G. (2020, July 24). 11 bed-ridden old age home members +ve, one died last month in Navi Mumbai. The Times of India. https://timesofindia.indiatimes.com/city/navi-mumbai/11-bed-ridden-old-age-home-members-ve-one-died-last-month-in-mumbai/articleshow/77138950.cms#:~:text=NAVI%20MUMBAI%3A%20As%20many%20as,succumbed%20to%20Covid%20during%20treatment

Padmanabha, U., Udayakiran, N., Nagarajaiah, P., & Kempaller, V. (2016). Morbidity profile of inmates in old age homes in Mangalore, South India. International Journal of Medical Science and Public Health, 5(11), 2230. https://doi.org/10.5455/ijmsph.2016.20032016461

Pandey, N., Tiwari, S., & Singh, I. (2012). Mental health problems among inhabitants of old age homes: A preliminary study. Indian Journal of Psychiatry, 54(2), 144. https://doi.org/10.4103/0019-5545.99533

Shaju Philip. (2020, August 2). Kerala: 95 COVID-19 cases in 2 old-age homes, state tightens norms. The New Indian Express. https://indianexpress.com/article/india/kerala/kerala-95-covid-19-cases-in-2-old-age-homes-state-tightens-norms-6534889/

Theresa, D. (2020, August 4). Care homes on high alert. The New Indian Express. http://cms.newindianexpress.com/cities/kochi/2020/aug/04/care-homes-on-high-alert-2178618.html

Van Bilsen, P., Hamers, J., Groot, W., & Spreeuwenberg, C. (2006). Demand of elderly people for residential care: An exploratory study. BMC Health Services Research, 6(1). https://doi.org/10.1186/1472-6963-6-39

Vijay Kumar, S. (2020, September 19). Coronavirus | 29 senior citizens recover from COVID-19 in Chennai’s Stanley Hospital. The Hindu. https://www.thehindu.com/news/cities/chennai/coronavirus-29-senior-citizens-recover-from-covid-19-in-chennais-stanley-hospital/article32644860.ece

World Health Organization. (2020, July 24). Older people and COVID-19. https://www.who.int/teams/social-determinants-of-health/covid-19

Zhang, Q., Li, M., & Wu, Y. (2020). Smart home for elderly care: Development and challenges in China. BMC Geriatrics, 20(1). https://doi.org/10.1186/s12877-020-01737-y

